# Digital intervention *mylovia* improves sexual functioning in women with sexual dysfunction in randomized controlled trial

**DOI:** 10.1101/2025.08.26.25334419

**Authors:** Wiebke Blaszcyk, Melanie Büttner, Linda T. Betz, Antje Riepenhausen, Gitta A. Jacob, Jan Philipp Klein, Johanna Schröder

**Author notes:** Correspondence to: Wiebke Blaszcyk, Research Department, GAIA, Hamburg, Germany.

## Abstract

Given the widespread issue of female sexual dysfunction and the scarcity of treatment options, novel therapeutic approaches are needed. This randomized controlled trial evaluated the use of *mylovia^©^*, a self-guided digital intervention for female sexual dysfunction and sexual pain disorder, in addition to treatment as usual (TAU) compared to TAU plus information material. 252 women participated. At three months, the intervention group showed significantly greater improvements (Cohen’s *d* = 0.51, *p* < 0.001) in sexual functioning, measured by the Female Sexual Function Index (FSFI), with effects maintained at six months. Clinical relevance was confirmed by Reliable Change Index (RCI) responder analysis. The intervention group also reported greater improvements in sexual desire, satisfaction, and pain-related cognitions and behaviors. There were no significant between-group differences in depressive symptoms or adverse events. The intervention demonstrated comparable efficacy to existing psychosocial treatments, offering a digital therapeutic that could narrow the current gender healthcare gap.

## Introduction

Impairments in sexual functioning are a common, yet often overlooked, health issue for women: according to a representative study, almost half of the sexually active women in Germany experienced at least one sexual problem within the past 12 months, while 17.5 % fulfilled the ICD-11 criteria for hypoactive sexual desire, sexual arousal dysfunction, orgasmic dysfunction, or sexual pain-penetration disorder^1^. To qualify for a clinical diagnosis, the sexual problem must occur frequently, although not necessarily always, over a period of at least several months, and be accompanied by clinically relevant distress^2^. For affected women, sexual dysfunction can severely impact other domains of physical and psychological wellbeing, such as quality of life^3,4^, self-esteem and body image^5^, as well as relationship quality^5,6^.

Despite the substantial health burden, female sexual dysfunctions often remain untreated: it is rarely addressed in outpatient psychotherapy settings^7,8^ or in primary care. In fact, within a German primary care sample, 84.8 % of women with clinically relevant levels of sexual dysfunction described themselves as untreated^9^. A major contributing factor is that female sexuality still carries stigma, shame and taboo, and is thus largely absent from clinical conversations^10–12^. Physicians, who are often sought out first by affected women, generally receive little formal training on sexuality^11–15^ and tend to either focus narrowly on isolated symptoms or refer to psychotherapists —who may themselves lack adequate expertise^15,16^. Consequently, many women are left without treatment and support. This gap is not simply an absence of evidence-based treatments, but the result of systemic gender bias: female sexual health has consistently been under-researched, undervalued, and overlooked in clinical settings^10,17–19^

This can be viewed as an example of gender health care inequity, given that male sexual dysfunctions have been much more extensively researched and addressed in the medical field. The blind spot for female sexual concerns in primary care assessments^11,20^, as well as the lack of clinical guidelines and shortage of treatments options, all showcase the differential attention placed on female versus male sexual problems. When female sexuality is considered at all, the emphasis often lies on reproduction or on enabling penetration rather than on promoting women’s sexual desire and pleasure^10,13,21–24^. This reveals the underlying heteronormativity, meaning the assumption of heterosexual relationships, gender norms, and sexual scripts privileging penile-vaginal intercourse as being normal and desirable^25^. Studies indicate that this, arguably male-centric, focus on vaginal intercourse does little to meaningfully support women’s sexual satisfaction^26–28^. All this is at odds with the importance placed on sexual health by the World Health Organization (WHO), which declares it to be “fundamental to the overall health and well-being of individuals, couples and families, and to the social and economic development of communities and countries” (WHO, https://www.who.int/health-topics/sexual-health).

In a departure from the traditionally narrow, heteronormative approach still seen in some research and clinical practice^25,29^, contemporary evidence-based psychotherapeutic treatments embrace a more comprehensive and biopsychosocial view of female sexuality—emphasizing desire, pleasure, and personal agency^10,30–33^. These approaches are largely grounded in cognitive-behavioral therapy (CBT) and typically include components such as (psycho-)education, body awareness, self-exploration, sensuality exercises, and techniques for enhancing (sexual) communication. Research supports the effectiveness of CBT-based interventions for sexual dysfunction^30,33^, although randomized controlled trials remain relatively scarce. Existing trials have found effect sizes around *d* = 0.5^34,35^.

Promisingly, digital interventions have also been shown to be effective in treating female sexual dysfunctions^34,36^. Given the scalability, ease of access and discreet nature of digital interventions, they might offer a valuable addition to the sexual health care landscape and help overcome identified treatment barriers such as stigma or lack of time and expertise^12^. This is particularly true for Germany, where digital therapeutics that have been vetted by the regulatory body (Federal Institute for Drugs and Medical Devices) are covered by the statutory health insurance and can be prescribed by physicians and psychotherapists, similarly to medication. One such digital therapeutic already exists for sexual pain-penetration disorder^21^, which is however less focused on other aspects of female sexuality and thus limited in scope. Currently, the German market lacks a digital therapeutic that comprehensively addresses all of the most common female sexual dysfunctions.

The present study evaluated the digital health application *mylovia*, which is based on cognitive-behavioral principles and centers female sexual pleasure. A randomized controlled trial (RCT) was conducted to assess the intervention’s effectiveness in improving sexual functioning in women with sexual dysfunction or sexual pain-penetration disorder. It was hypothesized that using the intervention in addition to treatment as usual (TAU) would lead to greater improvements in sexual functioning compared to TAU plus information material on treatment and counseling options. Empirically validated efficacy would make this intervention eligible for statutory health insurance coverage, thus providing a new treatment option for this underserved population.

## Results

### Participant flow and characteristics

Participants were recruited via an online campaign. 1,178 individuals expressed interest and were subsequently screened for eligibility, which led to 660 individuals being excluded, mostly due to incomplete data. This left 518 potential participants who were contacted for a diagnostic interview via telephone, of which 266 were excluded, mostly due to discontinued participation. Thus, 252 women were included in the study and randomized to either the intervention or control condition. This sample constituted the intention-to-treat (ITT) population, on which the analyses were based. 128 out of 129 intervention group participants (99.2%) activated the voucher needed to access *mylovia* and were therefore included in prespecified per protocol (PP) analyses, along with all control group participants, resulting in a PP population of 251 participants. Attrition was relatively low, with 11.6 % and 3.3 % of participants lost to follow-up at 3 months (T1) in the intervention and control group, respectively (Fig. 1).

**Fig. 1.**
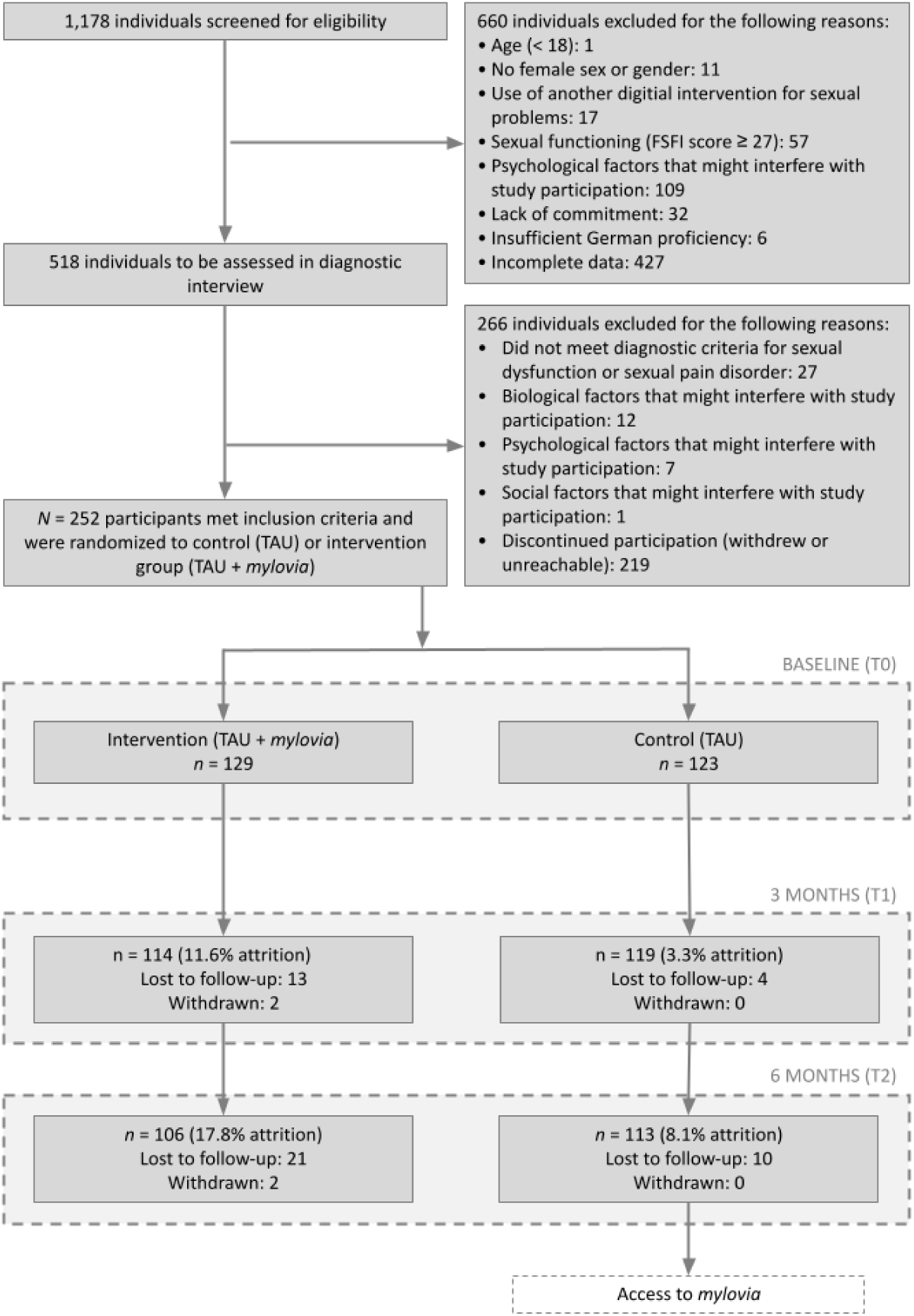
Participant flowchart.

Table 1 summarizes the main demographic and clinical characteristics of the study sample. The average participant age was 34.6 years. Educational attainment was relatively high, with about half of the participants (57.1 %) holding a university degree. Most participants were employed, either full-time (39.3 %) or part-time (32.9 %), and 88.9 % reported being in a romantic relationship. The majority identified as heterosexual (80.2 %) and ethnically White (95.6%). 84.1 % of participants were pre-menopausal and more than half (64.7 %) reported using some form of contraception.

**Table 1.** Baseline demographic and clinical characteristics.

| Variable | Control (n = 123) | Intervention (n = 129) |
| --- | --- | --- |
| **Age** (mean, SD) | 34.6 (9.5) | 34.5 (9.9) |
| **Education** [completed university studies], n (%) | 75 (61.0) | 69 (53.5) |
| **Employment** [part- or full time], n (%) | 90 (73.2) | 92 (71.3) |
| **Partnership situation** [single], n (%) | 15 (12.2) | 13 (10.1) |
| **Diagnosis** (*multiple possible*) |  |  |
| Sexual pain-penetration disorder, n (%) | 46 (37.4) | 41 (31.8) |
| Hypoactive sexual desire dysfunction n (%) | 70 (56.9) | 86 (66.7) |
| Sexual arousal dysfunction, n (%) | 37 (30.1) | 42 (32.6) |
| Anorgasmia, n (%) | 80 (65.0) | 66 (51.2) |
| **Sexual orientation** [heterosexual], n (%) | 99 (80.5) | 103 (79.8) |
| **Menopause status** [pre], n (%) | 103 (83.7) | 109 (84.5) |
| **In psychotherapy** [yes], n (%) | 26 (21.1) | 26 (20.2) |
| **Contraceptive method** [hormonal], n (%) | 32 (26.0) | 30 (23.3) |

Diagnostically, hypoactive sexual desire disorder was the most frequently reported issue (61.9 %), followed by anorgasmia (57.9 %) and sexual pain-penetration disorders (34.5 %). Using the Mini-DIPS diagnostic interview, major depressive disorder was found to be the most frequent psychiatric comorbidity, affecting roughly 19 % of participants. About one fifth (20.6 %) of the sample were in psychotherapy at the time of baseline assessment. As small percentage (13.9 %) of participants were taking psychotropic medication, most commonly antidepressants. Over the course of the clinical investigation, there was no statistically significant difference in relationship or treatment characteristics between the intervention and control groups (see Supplementary materials).

### Effectiveness

The primary endpoint to evaluate the effectiveness of *mylovia* was sexual functioning, assessed by the Female Sexual Function Index (FSFI), at three months (T1). The ITT analysis revealed that, after three months of using *mylovia* in addition to TAU, participants in the intervention group demonstrated significantly greater improvements in sexual functioning compared to those receiving TAU plus information material (estimated baseline-adjusted group difference = 3.7 points, 95% CI [1.7, 5.6], *p* < 0.001) with a Cohen’s *d* effect size of 0.51 (see Table 2 and Figure 2). This finding was corroborated by a conservative sensitivity analysis employing jump-to-reference (J2R) imputation, which estimated a group difference of 3.3 points (95% CI [1.6, 5.0], *p* < 0.001; *d* = 0.46). Similar results were observed in the prespecified per-protocol (PP) analysis (estimated baseline-adjusted group difference = 3.6 points, 95% CI [1.8, 5.5], *p* < 0.001; *d* = 0.51).

**Fig. 2.**
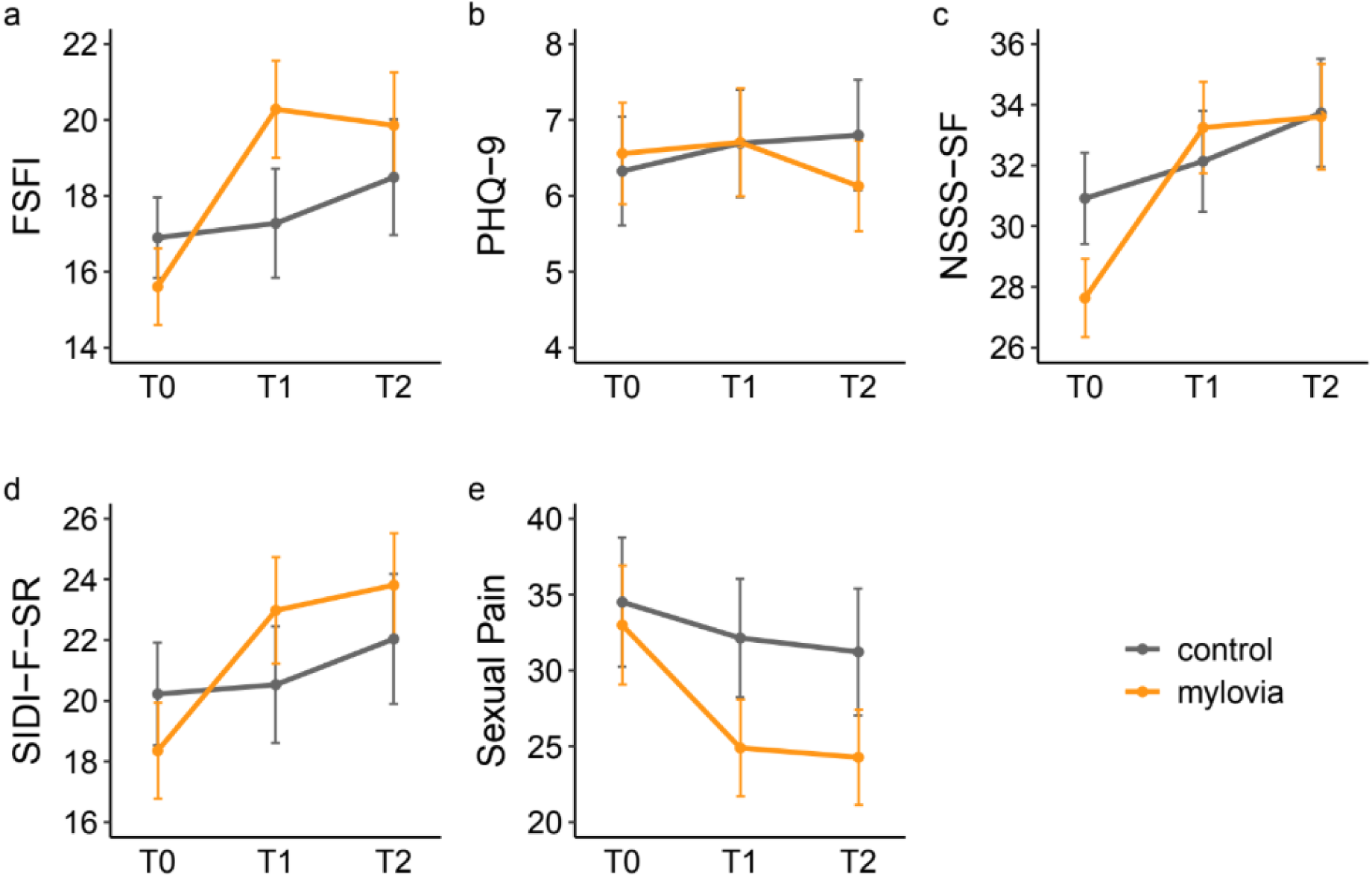
Mean scores for primary, secondary and exploratory endpoints for the *mylovia* group and control group across the study period (ITT analyses). a) Sexual Functioning (Female Sexual Function Index, FSFI, scale range: 2-36); b) Depressive Symptoms (Patient Health Questionnaire 9, PHQ-9, scale range: 0-27; c) Sexual Satisfaction (New Sexual Satisfaction Scale Short Form, NSSS-SF, scale range: 12-60); d) Sexual Desire (Sexual Interest and Desire Inventory-Female-Self-Report, SIDI-F-SR, scale range: 0-51); e) Cognitions and behaviors related to sexual pain (self-compiled questionnaire, scale range: 0-100). Error bars represent the 95% confidence interval (CI).

**Table 2.** Results of primary, secondary and exploratory endpoints (ITT analyses).

|  | Time | Control |  |  | *mylovia* |  |  | ANCOVA |  |  |  |
| --- | --- | --- | --- | --- | --- | --- | --- | --- | --- | --- | --- |
| | | n | mean | SD | n | mean | SD | Treatment effect (95% CI)<sup>a</sup> | *p* | Partial $\eta^2$ | *d* (95% CI)<sup>b</sup> |
| FSFI | T0 | 123 | 16.9 | 6.0 | 129 | 15.6 | 5.9 | - | - | - | - |
|  | T1 | 123 | 17.3 | 8.1 | 129 | 20.3 | 7.4 | 3.7 (1.7, 5.6) | < .001 | 0.06 | 0.51 (0.23, 0.79) |
|  | T2 | 123 | 18.5 | 8.6 | 129 | 19.9 | 8.1 | 2.0 (0, 4.0) | 0.048 | 0.02 | 0.26 (0, 0.52) |
| PHQ-9 | T0 | 123 | 6.3 | 4.1 | 129 | 6.6 | 3.9 | - | - | - | - |
|  | T1 | 123 | 6.7 | 4.0 | 129 | 6.7 | 4.1 | -0.1 (-0.9, 0.7) | 0.730 | 0 | 0.04 (-0.21, 0.30) |
|  | T2 | 123 | 6.8 | 4.1 | 129 | 6.1 | 3.5 | -0.8 (-1.6, 0) | 0.063 | 0.02 | 0.25 (-0.01, 0.5) |
| SIDI-F-SR | T0 | 123 | 20.2 | 9.6 | 129 | 18.4 | 9.1 | - | - | - | - |
|  | T1 | 123 | 20.5 | 10.9 | 129 | 23.0 | 10.2 | 3.1 (0.5, 5.6) | 0.020 | 0.03 | 0.30 (0.04, 0.56) |
|  | T2 | 123 | 22.0 | 12.1 | 129 | 23.8 | 9.9 | 2.2 (-0.6, 5.1) | 0.128 | 0.02 | 0.21 (-0.06, 0.48) |
| NSSS-SF | T0 | 123 | 30.9 | 8.5 | 129 | 27.6 | 7.5 | - | - | - | - |
|  | T1 | 123 | 32.1 | 9.4 | 129 | 33.2 | 8.7 | 2.6 (0.4, 4.8) | 0.020 | 0.03 | 0.31 (0.05, 0.58) |
|  | T2 | 123 | 33.7 | 10.1 | 129 | 33.6 | 10.1 | 1.6 (-0.9, 4.1) | 0.206 | 0.01 | 0.18 (-0.10, 0.45) |
| Sexual Pain | T0 | 123 | 34.5 | 24.1 | 129 | 33.0 | 22.7 | - | - | - | - |
|  | T1 | 123 | 32.1 | 22.1 | 129 | 24.9 | 18.5 | -6.2 (-9.6, -2.9) | < .001 | 0.06 | 0.49 (0.21, 0.76) |
|  | T2 | 123 | 31.2 | 23.6 | 129 | 24.3 | 18.2 | -5.8 (-9.5, -2) | 0.003 | 0.04 | 0.40 (0.14, 0.65) |

FSFI: Female Sexual Function Index, PHQ-9: Patient Health Questionnaire 9, SIDI-F-SR: Sexual Interest and Desire Inventory-Female-Self-Report, NSSS-SF: New Sexual Satisfaction Scale Short Form
<sup>a</sup> between-group difference on original scale 3 months after baseline, adjusted for baseline scores.
<sup>b</sup> based on baseline-adjusted means; positive values show effects in favor of the intervention group.

A responder analysis using the Reliable Change Index (RCI) criterion from baseline to T1 showed that a significantly higher proportion of participants in the intervention group (38.6 %) compared to the control group (16.0 %) experienced meaningful improvements in sexual functioning (χ² = 15.1, *p* < 0.001). This corresponds to an odds ratio of 3.3 (95% CI [1.8, 6.1]), indicating that the odds of reporting substantial improvement were 3.3 times higher in the *mylovia* group.

### Secondary and exploratory endpoints

At the six-month follow-up (T2), the ITT analysis indicated persistent beneficial effects of *mylovia*, with the intervention group showing a baseline-adjusted FSFI score 2.0 points higher than the control group (95% CI [0, 4.8], *p* = 0.048; *d* = 0.26).

There was no significant difference between groups for depressive symptoms (Patient Health Questionnaire 9, PHQ-9) with an estimated baseline-adjusted group difference of –0.1 points (95% CI = [-0.9, 0.7], *p* = 0.730; *d* = 0.04). At T2, the ITT analysis suggested a trend toward a between-group effect favoring the intervention group (estimated baseline-adjusted group difference = –0.8 points, 95% CI = [-1.6, 0], *p* = 0.063; *d* = 0.25).

To correct for multiple testing, a sequential gatekeeping strategy was prespecified and employed, in accordance with regulatory requirements. Given that no statistical difference was found for the first secondary outcome of depression, all subsequently planned endpoints – namely, the Sexual Interest and Desire Inventory-Female-Self-Report (SIDI-F-SR) and the New Sexual Satisfaction Scale Short Form (NSSS-SF) – were considered exploratory. The reporting of *p*-values is thus only for descriptive purposes.

After three months, participants in the *mylovia* group reported higher sexual desire (estimated baseline-adjusted difference on the SIDI-F-SR = 3.1 points, 95% CI [0.5, 5.6], *p* = 0.020; *d* = 0.30), greater sexual satisfaction (estimated baseline-adjusted group difference on the NSSS-SF = 2.6 points, 95% CI [0.4, 4.8], *p* = 0.020; *d* = 0.31), and significantly reduced maladaptive cognitions and behaviors related to sexual pain (estimated baseline-adjusted group difference on self-compiled questionnaire = –6.2 points, 95% CI [–9.6, –2.9], *p* < 0.001; *d* = 0.49). These findings were supported by both the J2R and PP analyses.

At T2, the intervention group continued to show higher scores in sexual desire and satisfaction, but these differences no longer reached statistical significance (estimated baseline-adjusted group difference on the SIDI-F-SR = 2.2 points, 95% CI = [-0.6, 5.1], *p* = 0.128; *d* = 0.21); estimated baseline-adjusted group difference on the NSSS-SF = 1.6 points, 95% CI = [-0.9, 4.1], *p* = 0.206; *d* = 0.18). However, improvements in sexual pain–related cognitions and behaviors remained statistically significant, with an estimated baseline-adjusted difference of –5.8 points (95% CI [–9.5, –2.0], *p* = 0.003; *d* = 0.40), indicating a sustained effect in this area. These findings were supported by both the J2R and PP analyses.

### Adverse effects

Adverse events, defined as unplanned medical treatments, were infrequent and occurred at a similar rate between the control and intervention groups at both T1 (4.2% vs. 2.7%, *p* = 0.519) and T2 (6.2% vs. 5.7%, *p* = 0.881). No adverse events were linked to the use of *mylovia*, and no adverse device effects were observed. Symptom worsening, measured by any decline in the total FSFI score, was significantly less common in the intervention group (21.1%) compared to the control group (42.0%) at T1 (χ² = 11.8, *p* < 0.001; OR = 0.37, 95% CI = [0.20, 0.66]). This difference in favor of the intervention group remained at T2 (24.5% vs. 32.7%), but was no longer statistically significant (χ² = 1.80, *p* = 0.180; OR = 0.67, 95% CI = [0.37, 1.21]). Overall, use of *mylovia* was less likely to worsen sexual functioning compared to TAU plus information material.

### Usage and user satisfaction

On average, users actively engaged with the program’s traceable features on 9.3 distinct days (SD = 5.8). However, it is worth noting that much of the program’s therapeutic engagement – including exercises, partner involvement, and personal exploration – is intended to occur offline and thus could not be tracked.

After three months, the intervention group rated their likelihood of recommending *mylovia* to a friend or colleague on average at 7.2 out of 10 (SD = 2.6), indicating overall satisfaction. However, participants noted that the recommendation question was unsuitable for the topic of sexual dysfunction, because they would not discuss this with friends or colleagues. This phrasing might have caused an underestimation of satisfaction.

Participants in the intervention group reported significantly greater subjective improvement in sexual functioning at T1, with a mean score of 4.7 (SD = 1.0) compared to 4.0 in the control group (SD = 1.1; t = –4.50, *p* < 0.001; *d* = 0.60, 95% CI = [0.33, 0.86]) on the Patient Global Impression of Change scale. Similarly, the intervention group reported a greater subjective improvement in their quality of life, scoring 4.5 (SD = 1.0) versus 4.2 for the control group (SD = 1.1; t = –2.34, *p* = 0.020; *d* = 0.31, 95% CI = [0.05, 0.57]).

### Subgroup analyses

Subgroup analyses based on participants’ baseline characteristics revealed that *mylovia* was effective regardless of psychotherapy use or method of contraception (Fig. 3). In the relatively small subgroup of peri– and postmenopausal women, the intervention effect was stable in size but not significant. Participants with a sexual pain-penetration disorder showed a smaller, non-significant effect (*d* = 0.30) compared to sexual dysfunctions (*d* = 0.65).

**Fig. 3.**
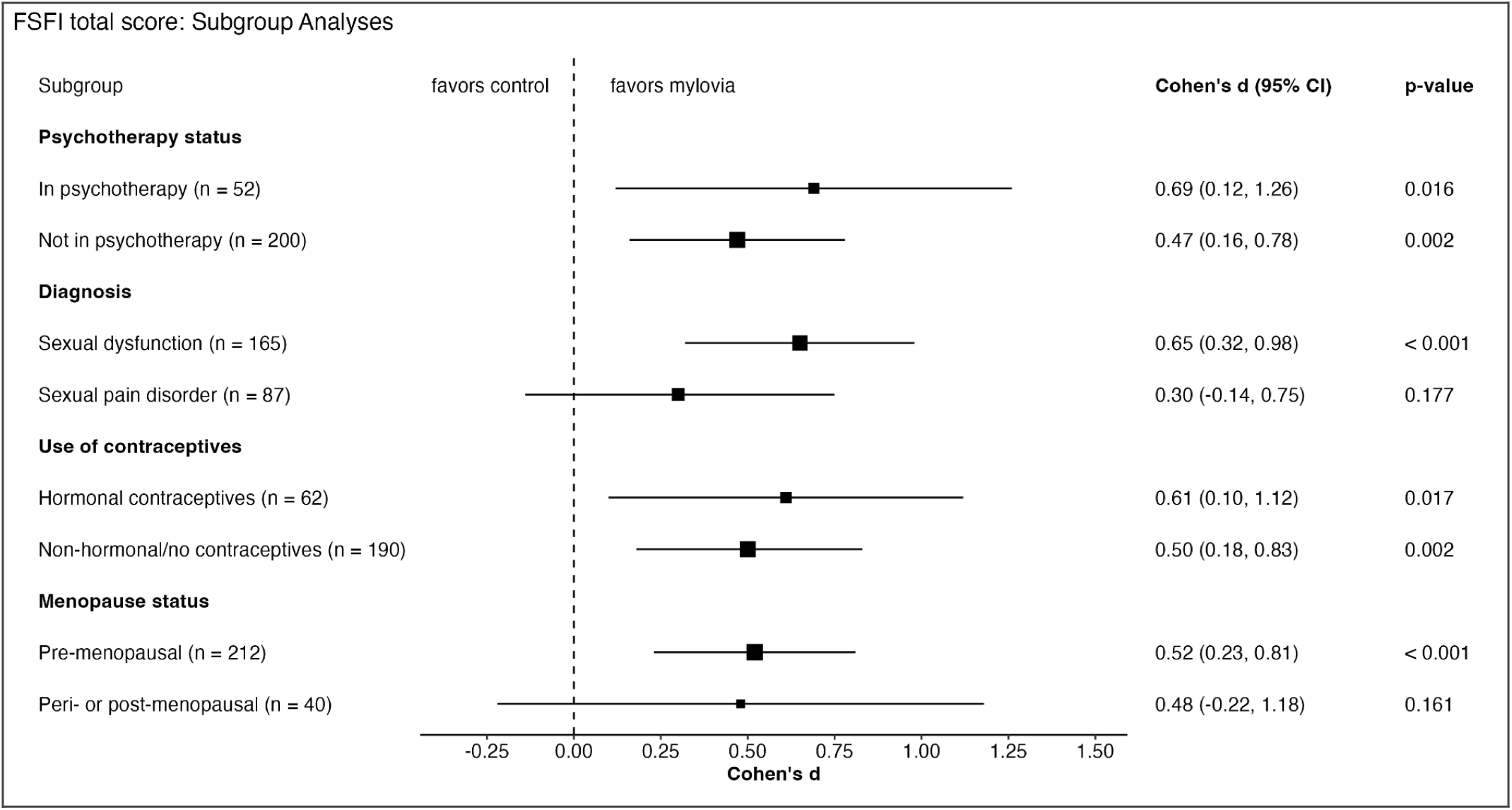
Forest plot of effect sizes (Cohen’s *d*) for the primary endpoint sexual functioning, assessed with the FSFI. *p*-values come from the ANCOVA. *Note*: Assigning multiple diagnoses was allowed. All participants with sexual pain-penetration disorder were assigned to the pain disorder subgroup, even if other diagnoses were also present.

## Discussion

In the present RCT with 252 participants, the online intervention *mylovia* was shown to be effective in increasing female sexual functioning within 3 months. Participants using *mylovia* in addition to TAU reported significant and, according to the RCI criterion, clinically relevant improvements in their sexual functioning compared to the control group, with a medium-sized between-group effect (*d* = 0.51; *p* < .001). The improvements were maintained at the 6-month follow-up. Users of *mylovia* also demonstrated significantly higher sexual desire and sexual satisfaction, as well as improved sexual pain-related cognitions and behaviors. These findings suggest that *mylovia* is an effective digital therapeutic, suitable for treating sexual dysfunction in women.

At follow-up, group differences were overall smaller and in the case of sexual desire and satisfaction no longer statistically significant. Utilization of additional outside support in the form of psychotherapy was comparable between groups at baseline, but increased more substantially in the control group over time – an imbalance that may help account for the smaller effect size observed at the six-month mark (see Table 1 and Supplementary Table 1).

Effectiveness was demonstrated regardless of whether participants were in psychotherapy at baseline and regardless of contraception method. For peri– and postmenopausal women, the intervention effect was stable but not significant, likely due to the subgroup’s small sample size. All subgroup results must be interpreted with caution because we did not perform a test for statistical moderation. Analyzing treatment effects across diagnostic subgroups suggested that individuals with sexual pain-penetration disorder experienced a more modest and statistically non-significant improvement (*d* = 0.30) compared to individuals with sexual dysfunction (*d* = 0.65). Although the reduced sample size of this subgroup might partially account for this, our finding also aligns with broader clinical research. For example, a meta-analysis on MBT interventions similarly reported a much lower treatment effect on sexual pain (*d* = 0.28) in comparison to other facets of sexual functioning^37^. Thus, sexual pain as a symptom domain might respond less to psychosocial interventions in general, not *mylovia* specifically, which may be linked to etiological factors: Sexual pain is often theorized to have, potentially unidentified, organic causes^38–40^. The presence of unaddressed or undiagnosed physiological factors could therefore explain *mylovia*’s more limited impact on pain. Furthermore, sexual pain-penetration disorder is frequently characterized by greater severity^1^, which is corroborated by the significantly lower FSFI scores this subgroup exhibited at baseline.

No significant effects were found for the secondary outcome of depressive symptoms, which is likely explained by the sample’s, on average, lack of clinically relevant levels of depression at baseline, indicated by a low PHQ-9 mean score, and thus a possible floor effect. This can also be viewed as a strength of the study, since it implies that the increased sexual functioning is not merely the result of reduced depression.

Overall, our findings are in line with previous research. *mylovia*’s therapeutic framework is largely based on Mindfulness-Based Therapy (MBT). MBT, a subtype of Cognitive Behavioral Therapy (CBT), has in recent years gained attention and evidence as a treatment for female sexual dysfunction. A 2017 meta-analysis showed MBT’s efficacy for female sexual functioning varied by domain, with effect sizes from *d* = 0.28 (pain) to *d* = 0.63 (sexual arousal)^37^. Another meta-analysis found a moderate-sized effect (*d* = 0.55) on female and male sexual functioning^41^. Except for one, all interventions included in these meta-analyses were carried out in-person. Thus, the current results extend previous findings by showing that a digital intervention can achieve similar effects.

More traditional CBT has also shown efficacy for female sexual dysfunction^30^, but supporting empirical data is limited in quantity and quality. Typical CBT approaches in the treatment of sexual dysfunction include psychoeducation, cognitive restructuring, sensuality exercises/sensate focus, guided masturbation, desensitization/exposure therapy, and communication strategies^30^. Randomized controlled trials and meta-analytic data on CBT for female sexual dysfunction are scarce. A recent meta-analysis reported a large effect (d = 1.34) for face-to-face CBT on overall sexual functioning, but this was based on only three RCTs of generally low quality and with very small sample sizes. In contrast, a much smaller effect (d = 0.58) was reported in a 2013 meta-analysis looking at various, predominantly in-person, psychological interventions (including CBT)^35^. Similarly, a 2022 systematic review and meta-analysis of CBT-based online interventions found a medium effect of *g* = 0.59 on female sexual functioning^34^. Thus, overall, the observed effect size for *mylovia* (*d* = 0.51) is comparable to previously studied MBT and CBT interventions, including both online and face-to-face delivery. Notably, *mylovia* was able to achieve this effect as a fully self-guided program whereas most digital interventions in the meta-analysis provided varying degrees of human guidance.

This RCT benefits from a large sample size, which is rare in this field, and low dropout rates compared to similar trials where attrition in the intervention group ranged from 22% to 42% after 3 months^21,42^. This, together with the pragmatic study design, increases the generalizability of our results. We implemented a comprehensive outcome assessment by including endpoints like sexual desire, satisfaction, and pain-related cognitions and behaviors, providing a holistic picture of the intervention’s impact. Additionally, the use of responder analyses and as J2R sensitivity analyses support the robustness of our findings. Future research might include examining the effectiveness for trans and non-binary populations, as well as exploring specific mechanisms of change.

This study also comes with some limitations. Our sample was self-selected and thus might have favored women of a certain educational level and technological affinity – although this does arguably correspond to the future user population. Further, there was no blindness to allocation for participants, which may have inflated the effect. However, for this type of intervention, it is virtually impossible to create an active control condition without therapeutic content but still convincing to participants^43^. The pragmatic comparison to TAU is thus standard practice in trials like this.

Overall, this study addresses an unmet need and a substantial gap in evidence-based treatment options for female sexual dysfunction. The fully self-guided program *mylovia* demonstrated comparable efficacy to existing psychosocial interventions for female sexual dysfunction and sexual pain, with added digital benefits like discretion, accessibility, and lower cost. This strongly suggests that digital therapeutics like *mylovia* should form part of the standard treatment approach, and future research should explore how they can best be deployed to improve female sexual healthcare, thereby leveraging their potential to narrow the gender healthcare gap.

## Methods

### Study design

This pragmatic, randomized controlled study investigated the efficacy of the digital intervention *mylovia* when used in addition to TAU versus TAU plus information material in adult women with sexual dysfunction or sexual pain-penetration disorder in Germany (ClinicalTrials.gov ID: NCT06237166, https://clinicaltrials.gov/study/NCT06237166, date of registration: 24 January, 2024). The ethics committee of the Hamburg State Chamber of Physicians (Ärztekammer Hamburg) reviewed and approved the clinical investigation protocol (reference number 2024-101233-BO-ff), and the trial adhered to the Declaration of Helsinki. All participants received comprehensive information and provided informed consent before joining the study. All assessments and other contact with the participants occurred online or via telephone.

### Participants and procedure

Participants were recruited through online advertisements on Google and Meta platforms. Enrollment began in May 2024, with the final follow-up data collected in May 2025. Potential participants accessed the study website for trial details and registered interest via a contact form. They then received a link to a secure survey tool (*LimeSurvey*) for participant information, consent, and data protection details. After signing the consent form, they completed an online survey for inclusion and exclusion criteria, demographics, and baseline data. *LimeSurvey* was used to collect all assessment data at baseline (T0), 3 months (T1) and 6 months (T2). As an incentive, participants received a 10€ gift card per completion of the 3-month (T1) and 6-month (T2) assessments, so up to 20€ total.

To be eligible, participants had to be at least 18 years old, be of female sex and gender, possess their own smartphone or computer for internet access, have sufficient German language comprehension, and consent to participate in the study. They further had to score below 27 on the FSFI – the cut-off point indicating sexual dysfunction^44^ – and fulfill diagnostic criteria for sexual dysfunction (ICD-11: HA00, HA01, HA02) or sexual pain-penetration disorder (ICD-11: HA20), which was verified by trained study personnel via the DISEX-F interview^45^.

As sexual dysfunction is a biopsychosocial phenomenon, the intervention was hypothesized to benefit women regardless of etiology (biological, psychological, or social). However, following assessment, individuals were excluded if one of these domains was severely compromised, meaning if biological (e.g., cancer, multiple sclerosis), psychological (e.g., schizophrenia, bipolar disorder, borderline personality disorder, severe depression, acute suicidality, substance use disorder) or social (e.g. severe partnership problems, domestic violence) factors were identified that would likely have interfered substantially with study participation. Use of another digital intervention for sexual problems was also an exclusion criterion.

Following confirmation of eligibility, participants were randomly allocated in a 1:1 ratio to one of two study arms: the intervention group (*mylovia* + TAU) or the control group (TAU + information material). Randomization followed a simple randomization procedure implemented via a computer-generated sequence in an automated system (akin to a digital coin toss) integrated into the study platform. Allocation was triggered directly upon eligibility verification and was concealed from study personnel. Due to the trial’s pragmatic nature, participants were aware of their assigned group. As all outcome data were obtained through self-report instruments, there were no independent outcome assessors, and blinding at the assessment level was not applicable. Participants in the intervention group received immediate 180-day access to *mylovia*, whereas participants in the control group received a two-page informational signposting document on treatment and counseling options, and were offered access to *mylovia* after they had completed their final assessment at T2.

### Intervention

*mylovia* is a self-guided online intervention, developed by a multidisciplinary team headed by GAIA (Hamburg, Germany), and is aimed at improving women’s sexual functioning. It is accessible via any web-enabled device. The interactive program simulates therapist dialogues through predefined text-based exchanges, adapting content and offering tailored exercises based on user responses (see Fig. 4). It also offers audio recordings and PDF materials. Users regularly receive motivational messages and reminders via email or text message to encourage engagement with the program or implementation of exercises and homework.

**Fig. 4.**
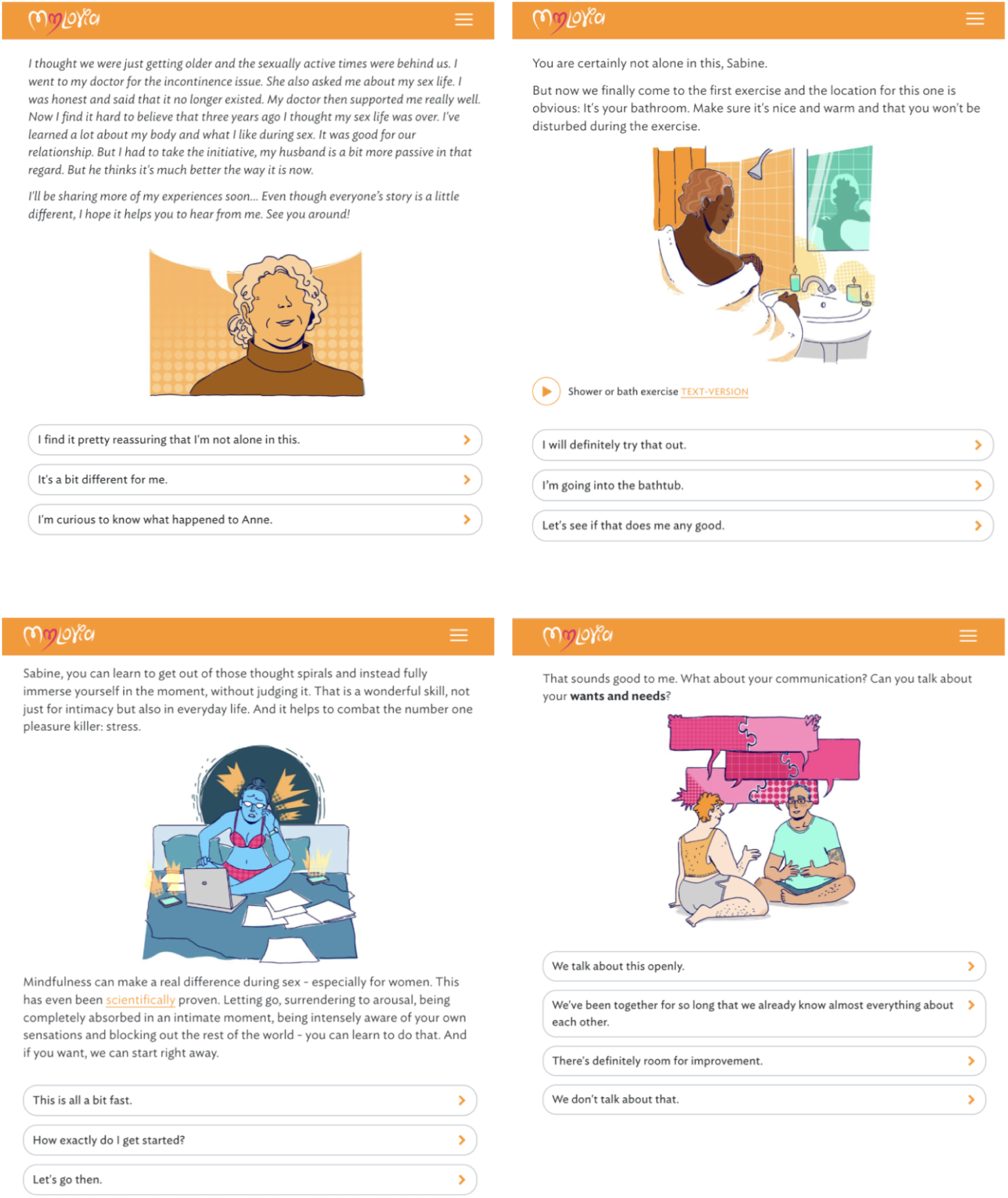
Selected screenshots of *mylovia*, translated to English for illustration purposes. *Note:* The actual *mylovia* digital therapeutic is in German. An English version is currently not available. (please see separate pdf for higher resolution)

The therapeutic content is based on CBT principles and also draws extensively on mindfulness-based approaches to female sexual dsyfunction^37,46^, covering several therapeutic areas (Table 3).

**Table 3.** Therapeutic areas covered by the *mylovia* digital intervention.

|  |  |
| --- | --- |
| **Sexual knowledge** | The program provides comprehensive psychoeducation and sex education, correcting common myths and offering fundamental facts about anatomy and sexual health to foster healthier sexual scripts. |
| **Relaxation and mindfulness** | Rooted in mindfulness-based sex therapy, *mylovia* enhances body awareness and sensitivity through mindful self-observation and non-judgmental present-moment focus, alongside relaxation and stress management techniques. |
| **Sexual skills** | Users engage in diverse exercises to expand sexual skills, including self-exploration, mindful masturbation, pelvic floor exercises, and sensate focus, with guidance for practical implementation. |
| **Sexual desire** | *mylovia* helps users explore sexual fantasies and arousal sources, reframing desire as an incentive-motivation system, and provides exercises to integrate more sensuality into daily life. |
| **Sexual pain** | Users who suffer from pain-penetration disorder, receive education, and are guided through cognitive defusion, pelvic floor relaxation, and dilator training. |
| **Body image and self-esteem** | The program encourages critical reflection on beauty ideals and body image, offering exercises like guided mirror exposure to foster positive self-perception and boost self-esteem. |
| **Partnered sex** | *mylovia* helps users address unrealistic partnered sex ideals and communication challenges, providing guidance for open, constructive dialogue, including sharing preferences and setting boundaries. |

### Outcomes and assessments

The primary endpoint of sexual functioning was assessed using the FSFI^47,48^, a well-established 19-item self-report measure covering six domains of sexual function in women, namely desire, arousal, lubrication, orgasm, satisfaction, and pain. The internal consistency at baseline was Cronbach’s α = 0.93.

For the planned gatekeeping testing strategy (see Statistical analysis), the secondary outcomes were prioritized in the following order: depressive symptoms, measured by the Patient Health Questionnaire-9 (PHQ-9, α = 0.80)^49,50^; sexual desire measured by the Sexual Interest and Desire Inventory Female Self-Report (SIDI-F-SR, α = 0.87)^51,52^; and sexual satisfaction, measured by the New Sexual Satisfaction Scale Short Form (NSS-SF, α = 0.81)^53,54^. Additionally, a self-compiled questionnaire assessing cognitions and behaviors related to sexual pain was used as an exploratory endpoint (α = 0.84).

At baseline, various sociodemographic (e.g. age, partnership status, sexual orientation, ethnicity) and clinical (e.g. psychotherapy use, history of sexual abuse, contraception) were assessed. The Mini-DIPS^55^ diagnostic interview was used to screen for psychiatric comorbidities.

User satisfaction was evaluated by asking participants to rate their likelihood of recommending *mylovia* to a friend or colleague, ranging from 0 to 10. Additionally, the Patient Global Impression of Change scale^56^ served to measure the subjective improvement in sexual functioning and quality of life. All assessments were carried out in German.

### Safety and adverse events

Adverse events, operationalized as unplanned outpatient and inpatient treatments, were monitored throughout the study. Participants were asked to report any adverse events at T1 and T2 and were also able to report them directly to the study team via email, phone, or website contact form.

### Sample Size

The required sample size was determined via a priori power analysis using the R package *pwr*^57^. To account for the broad inclusion criteria designed to capture a diverse population, we assumed a slightly more conservative effect size of *d* = 0.40 than observed in previous studies^21,34,35^. To detect this effect with 80% power at α = .05 (two-sided), 100 participants per group were required. Allowing for 20% dropout, the final target sample was set at 250 (2 × 125).

### Statistical analysis

Treatment effects were estimated as baseline-adjusted mean differences with 95% CIs, based on ANCOVA models. In these models, the outcome at follow-up (3 or 6 months) served as the dependent variable, treatment condition (intervention vs. control) as the independent variable, and the corresponding baseline value of the outcome as the covariate. Standardized effect sizes (Cohen’s *d*) were derived from estimated marginal means using the *emmeans* package^58^.

The primary analysis followed the intention-to-treat (ITT) principle, including all randomized participants. Missing data were handled under a missing-at-random assumption with bootstrapped multiple imputation, using the R packages *bootImpute* and *mice*^59,60^.

Specifically, 1,000 bootstrap samples of the observed data were generated using *bootImpute*. Each sample was then, as recommended, imputed twice using *mice* with predictive mean matching. Imputation models included baseline scores, group allocation, and additional sociodemographic and clinical variables (age, psychotherapy at baseline, diagnosis of sexual pain-penetration disorder, method of contraception, menopause status and intake of any psychotropic medication at baseline). Prespecified subgroup analyses within the ITT population were conducted for psychotherapy status, diagnosis, contraception method, and menopause status, based on ANCOVA models.

In parallel, we conducted a per-protocol (PP) analysis, which applied the same statistical procedures as the primary analysis, but included only those participants from the intervention group who had activated their access to *mylovia*, as well as all control group participants. To assess robustness to the missing data mechanism, we conducted a conservative sensitivity analysis using jump-to-reference (J2R) with the R packages *bootImpute* and *mlmi*^59,61^. This approach assumes that participants discontinuing the intervention follow the outcome trajectory of the control group from the point of dropout onward ^62^.

Statistical significance was defined as *p* < .05 (two-sided). A hierarchical gatekeeping strategy was applied to control for multiple comparisons across secondary outcomes, with a prespecified testing order (see Outcomes and assessments). To evaluate clinical relevance, a responder analysis was conducted for the primary outcome using the Reliable Change Index (RCI)^63^. Participants with RCI scores > 1.96 from baseline to 3-month follow-up were classified as responders. Group differences in responder rates were tested using χ² tests and expressed as odds ratios (OR). All analyses were performed with *R*, version 4.4.1. The analyses were pre-defined in the study protocol, which was finalized before data access for analysis.

## Data availability

Due to proprietary restrictions, the datasets for this study are not publicly accessible. However, they can be obtained from the corresponding author upon reasonable request.

## Code availability

Not applicable (no proprietary code was used in data collection or analysis).

## Acknowledgements

This trial was funded by GAIA (Hamburg, Germany). The authors thank all the individuals who participated in the *mylovia* trial.

## Author contributions

GAJ and AR designed the study with substantial input from JS. GAJ contributed to the development of the intervention tool with substantial input from MB. WB contributed to and supervised the study operations. WB, LTB and GAJ collected the data. LTB formally analyzed the data with substantial input from WB, JPK and JS. WB and LTB interpreted the data, with substantial input from JS. WB wrote the original draft and final manuscript, with substantial input from all authors. All authors critically revised the manuscript and approved the final version.

## Competing interests

WB, AR, LTB and GAJ are employed by GAIA, the developer and manufacturer of the intervention described in the study. JPK received funding for clinical trials (German Federal Ministry of Health, Servier), payments for presentations on psychological internet interventions (GAIA, Oberberg, Servier, Stillachhaus), consulting fees from developers and distributors of psychological internet interventions (all about me, Boehringer, Ethypharm, GAIA, medac, sympatient) payments for workshops and books (Beltz, Elsevier, Hogrefe and Springer) on psychiatry, psychosomatics and psychotherapy. He is past president of the CBASP network (now DsG-CBASP) and serves as vice chairman of the chapters “Digital Psychiatry” and “Psychotherapy” of the German Psychiatric Association (DGPPN). JS and MB have no conflicts of interest to declare.

## Additional information

**Supplementary information** Supplementary information is available.

**Correspondence** and requests for materials should be addressed to Wiebke Blaszcyk.

## Notes

### Clinical Trial

NCT06237166

### Author Declarations

The ethics committee of the Hamburg State Chamber of Physicians (Ärztekammer Hamburg) gave ethical approval for this work.

